# Are commercial antibody assays substantially underestimating SARS-CoV-2 ever infection? An analysis on a population-based sample in a high exposure setting

**DOI:** 10.1101/2020.12.14.20248163

**Authors:** Gheyath K. Nasrallah, Soha R. Dargham, Farah Shurrab, Duaa W. Al-Sadeq, Hadeel Al-Jighefee, Hiam Chemaitelly, Zaina Al Kanaani, Abdullatif Al Khal, Einas Al Kuwari, Peter Coyle, Andrew Jeremijenko, Anvar Hassan Kaleeckal, Ali Nizar Latif, Riyazuddin Mohammad Shaik, Hanan F. Abdul Rahim, Hadi M. Yassine, Mohamed G. Al Kuwari, Hamda Qotba, Hamad Eid Al Romaihi, Patrick Tang, Roberto Bertollini, Mohamed Al-Thani, Asmaa A. Althani, Laith J. Abu-Raddad

**Affiliations:** Biomedical Research Center, Member of QU Health, Qatar University, Doha 2713, Qatar; Department of Biomedical Science, College of Health Sciences, Member of QU Health, Qatar University, Doha 2713, Qatar; Infectious Disease Epidemiology Group, Weill Cornell Medicine - Qatar, Cornell University, Qatar Foundation - Education City, Doha, Qatar; World Health Organization Collaborating Centre for Disease Epidemiology Analytics on HIV/AIDS, Sexually Transmitted Infections, and Viral Hepatitis, Weill Cornell Medicine – Qatar, Cornell University, Qatar Foundation – Education City, Doha, Qatar; Hamad Medical Corporation, Doha, Qatar; College of Health Sciences, QU Health, Qatar University, Doha, Qatar; Primary Health Care Corporation, Doha, Qatar; Ministry of Public Health, Doha, Qatar; Department of Pathology, Sidra Medicine, Doha, Qatar; Department of Population Health Sciences, Weill Cornell Medicine, Cornell University, New York, USA

**Keywords:** COVID-19, coronavirus, serology, antibodies, PCR, Qatar

## Abstract

**Background:** Performance of three automated commercial serological IgG-based assays was investigated for assessing SARS-CoV-2 ever (past or current) infection in a population-based sample in a high exposure setting.

**Methods:** PCR and serological testing was performed on 394 individuals.

**Results:** SARS-CoV-2-IgG seroprevalence was 42.9% (95% CI 38.1%-47.8%), 40.6% (95% CI 35.9%-45.5%), and 42.4% (95% CI 37.6%-47.3%) using the CL-900i, VidasIII, and Elecsys assays, respectively. Between the three assays, overall, positive, and negative percent agreements ranged between 93.2%-95.7%, 89.3%-92.8%, and 93.8%-97.8%, respectively; Cohen kappa statistic ranged from 0.86-0.91; and 35 specimens (8.9%) showed discordant results. Among all individuals, 12.5% (95% CI 9.6%-16.1%) had current infection, as assessed by PCR. Of these, only 34.7% (95% CI 22.9%-48.7%) were seropositive by at least one assay. A total of 216 individuals (54.8%; 95% CI 49.9%-59.7%) had evidence of ever infection using antibody testing and/or PCR during or prior to this study. Of these, only 78.2%, 74.1%, and 77.3% were seropositive in the CL-900i, VidasIII, and Elecsys assays, respectively.

**Conclusions:** All three assays had comparable performance and excellent agreement, but missed at least 20% of individuals with past or current infection. Commercial antibody assays can substantially underestimate ever infection, more so when infection rates are high.

## Introduction

Coronavirus disease 2019 (COVID-19), due to the novel severe acute respiratory syndrome coronavirus 2 (SARS-CoV-2), continues to be a global health challenge. As of November 22, 2020, the COVID-19 burden included 57.6 million confirmed cases and 1.3 million deaths worldwide [1]. Meanwhile, the true extent of exposure to the SARS-CoV-2 infection and how far different national populations are from herd immunity remain poorly understood. Commercial serological assays are increasingly being used to address this gap in evidence. The extent to which such assays can capture ever infection in a population remains to be elucidated.

Understanding who has been exposed and potentially acquired immunity against this virus may help healthcare providers and public health stakeholders in establishing and implementing more efficient and effective strategies and policies for managing the disease and economic burden associated with the COVID-19 pandemic.

Qatar experienced a large SARS-CoV-2 epidemic with a high rate of laboratory-confirmed infections at >60,000 infections per million population [2-4]. As part of the national response, the public health authorities expanded serological testing for SARS-CoV-2 antibodies for both healthcare and research purposes. Three automated main serological testing platforms are being used. The first is the Roche Elecsys® Anti SARS CoV 2 (Roche, Switzerland) [5] platform at Hamad Medical Corporation (HMC), the main public healthcare provider and the nationally-designated provider for all COVID-19 healthcare needs. The second is the Mindray CL-900i anti-SARS-CoV-2 IgG (Shenzhen Mindray Bio-Medical Electronics Co., China) [6] platform at Qatar University (QU), which is used for research purposes. The third is the BioMérieux VidasIII (BioMérieux, Marcy-l’Etoile, France) [7] platform at QU, which is also being used for research purposes.

To interpret the emerging results of serological testing and to inform the national response, this study was conducted to compare the performance of these three assays and to assess the implications for measuring SARS-CoV-2 ever infection. The novelty and strength of this study is that it is conducted based on a population-based sample [8] in a setting at a high exposure to this infection [2, 3, 9, 10].

## Methods

Blood specimens were collected from 394 volunteering individuals between July 26 and September 9, 2020, as a sub-study of a nationwide survey [8] assessing SARS-CoV-2 seroprevalence (IgG antibodies) and current-infection prevalence (using polymerase chain reaction [PCR] testing) in the wider population of craft and manual workers who constitute 60% of the population of Qatar [11]. Informed by prior work [12, 13], a sample size of 400 was estimated to be sufficient to ensure narrow confidence intervals for the Cohen’s kappa statistic, but we were able to include and test only 394 specimens. The research work was approved by the ethics review boards at HMC, QU, and Weill Cornell Medicine-Qatar.

The automated serological testing was performed using the above indicated three commercial assays. The Roche Elecsys® Anti SARS-CoV-2 (“Elecsys” in short form) assay, our reference assasy, uses a recombinant protein representing the nucleocapsid (N) antigen for the determination of IgG antibodies against SARS-CoV-2 [5]. Anti-SARS-CoV-2 results were generated following the manufacturer’s instructions (reactive: optical-density cutoff index ≥1.0 vs. non-reactive: cutoff index <1.0) [5, 14].

The Mindray CL-900i® anti-SARS-CoV-2 IgG (“CL-900i” in short form) assay uses paramagnetic microplates coated with recombinant nucleocapsid (N) and spike (S) antigens for the determination of anti-SARS-CoV-2 IgG antibodies [6]. The analyzer automatically calculates the analyte concentration of each serum specimen according to a master calibration curve, and the results are shown in the units of U/mL. Anti-SARS-CoV-2 results were generated following the manufacturer’s instructions (reactive: optical-density cutoff index ≥10.0 vs. non-reactive: cutoff index <10.0) [6, 15].

The BioMérieux VidasIII assay (“VidasIII” in short form) uses a VIDASIII® analyzer for anti-SARS-CoV-2 IgG detection through a two-step sandwich ELFA assay [7]. The IgG in the serum specimen binds to a recombinant spike S1 sub-domain (containing the receptor-binding domain [S1-RBD]) of the SARS-CoV-2 virus coated on a solid phase. Alkaline phosphatase-conjugated anti-human IgG are then added. The fluorescence intensity generated by the substrate is then measured at a wavelength of 450 nm. The intensity of the signal is proportional to the level of IgG. The optical-density cutoff index was calculated according to the manufacturer’s instructions [7, 14]. The ratio between the relative fluorescence value (RFV) measured in the specimen and the RFV from the calibrator was interpreted as positive if the index value was ≥1.0 [7, 14].

All PCR testing was conducted at HMC Central Laboratory or at Sidra Medicine Laboratory, following standardized protocols. Nasopharyngeal and oropharyngeal swabs (Huachenyang Technology, China) were collected and placed in Universal Transport Medium (UTM). Aliquots of UTM were: extracted on the QIAsymphony platform (QIAGEN, USA) and tested with real-time reverse-transcription PCR (RT-qPCR) using the TaqPath™ COVID-19 Combo Kit (Thermo Fisher Scientific, USA) on an ABI 7500 FAST (ThermoFisher, USA); extracted using a custom protocol [16] on a Hamilton Microlab STAR (Hamilton, USA) and tested using the AccuPower SARS-CoV-2 Real-Time RT-PCR Kit (Bioneer, Korea) on an ABI 7500 FAST; or loaded directly to a Roche cobas® 6800 system and assayed with the cobas® SARS-CoV-2 Test (Roche, Switzerland). The first assay targets the S, N, and ORF1ab regions of the virus; the second targets the virus’ RdRp and E-gene regions; and the third targets the ORF1ab and E-gene regions.

Results of the serological and PCR testing were subsequently linked to the HMC centralized and standardized database comprising all SARS-CoV-2 PCR testing conducted in Qatar since the start of the epidemic [2, 17]. The database also includes data on hospitalization and on the World Health Organization (WHO) severity classification [18] for the hospitalized PCR-confirmed infections.

Results from the three types of serological testing were cross-tabulated. Four concordance metrics were estimated: overall, positive, and negative percent agreement, as well as Cohen’s kappa statistic. The latter is a robust metric that measures the level of agreement, beyond chance, between two diagnostic testing methods [19]. The kappa statistic ranges between 0 and 1; a value ≤0.40 indicates poor agreement, a value between 0.40 and 0.75 indicates fair/good agreement, and a value ≥0.75 indicates excellent agreement [19]. Level of significance was established at 5%, and a 95% confidence interval (CI) was reported for each metric. A nonparametric statistical method, Spearman correlation, was used to assess the correlation between the optical densities of each pair of antibody assays. Calculations were conducted using Microsoft Excel.

## Results

SARS-CoV-2 seroprevalence was estimated at 42.9% (169/394; 95% CI 38.1%-47.8%) using the CL-900i assay, 40.6% (160/394; 95% CI 35.9%-45.5%) using the VidasIII assay, and 42.4% (167/394; 95% CI 37.6%-47.3%) using the Elecsys assay. A total of 183 specimens were seropositive in at least one of the assays for a total sample seroprevalence of 46.5% (183/394; 95% CI 41.6%-51.4%).

Table S1 shows the results of the serological and PCR testing for each of the 394 participants. A total of 35 specimens showed discordant results between the three antibody assays (Table 1). Of the 35 individuals with discordant antibody results, 9 were PCR-positive at the time of specimen collection. Eleven specimens were seropositive using the CL-900i assay but seronegative using the VidasIII and the Elecsys assays; among these, two were PCR-positive with cycle threshold (Ct) values of 23.9 and 27.0. Five specimens were seropositive using the Elecsys assay but seronegative using the VidasIII and the CL-900i assays; among these, one person was PCR-positive with a Ct value of 21.6. Two specimens were seropositive using the VidasIII assay but seronegative using the CL-900i and the Elecsys assays; among these, one was PCR-positive with a Ct value of 29.2.

**Table 1.** Characteristics of the specimens that were discordant between the Mindray CL-900i anti-SARS-CoV-2 IgG, BioMérieux VidasIII anti-SARS-CoV-2 IgG, and Roche Elecsys Anti SARS-CoV-2 antibody testing. There were no PCR-confirmed infections *prior* to the study for any of the persons listed in this table.

| Sample number<sup>%</sup> | Mindray CL-900i anti-SARS-CoV-2 IgG<sup>*</sup> | | BioMérieux VidasIII anti-SARS-CoV-2 IgG<sup>**</sup> | | Roche Elecsys Anti SARS-CoV-2<sup>***</sup> | | PCR test at time of study | | Symptomatic at time of study<sup>#</sup> | Severity at or right after time of study<sup>\$</sup> |
| --- | --- | --- | --- | --- | --- | --- | --- | --- | --- | --- |
|  | Optical density | Test Result | Optical density | Test Result | Optical density | Test Result | Test result | Ct value |  |  |
| 20 | 3.76 | Negative | 5.86 | Positive | 0.48 | Negative | Negative |  | No | N/A |
| 21 | 111.72 | Positive | 0.45 | Negative | 67.27 | Positive | Negative |  | No | N/A |
| 22 | 3.74 | Negative | 0.10 | Negative | 8.40 | Positive | Positive | 21.62 | Symptomatic | Severe |
| 29 | 5.81 | Negative | 1.08 | Positive | 5.62 | Positive | Negative |  | No | N/A |
| 37 | 7.77 | Negative | 0.47 | Negative | 3.66 | Positive | Negative |  | No | N/A |
| 40 | 20.24 | Positive | 0.01 | Negative | 0.16 | Negative | Negative |  | No | N/A |
| 50 | 31.25 | Positive | 0.04 | Negative | 0.09 | Negative | Negative |  | No | N/A |
| 61 | 135.05 | Positive | 0.45 | Negative | 38.01 | Positive | Negative |  | No | N/A |
| 71 | 4.17 | Negative | 0.88 | Negative | 3.01 | Positive | Negative |  | No | N/A |
| 82 | 71.76 | Positive | 0.02 | Negative | 0.08 | Negative | Negative |  | No | N/A |
| 126 | 10.50 | Positive | 0.15 | Negative | 0.18 | Negative | Positive | 26.95 | No | N/A |
| 128 | 220.63 | Positive | 0.62 | Negative | 7.72 | Positive | Positive | 33.81 | No | N/A |
| 129 | 16.60 | Positive | 0.07 | Negative | 0.08 | Negative | Negative |  | No | N/A |
| 131 | 36.22 | Positive | 0.46 | Negative | 9.23 | Positive | Positive | 28.32 | No | N/A |
| 137 | 4.72 | Negative | 0.46 | Negative | 10.98 | Positive | Negative |  | No | N/A |
| 147 | 39.57 | Positive | 0.94 | Negative | 10.92 | Positive | Negative |  | No | N/A |
| 174 | 12.05 | Positive | 1.29 | Positive | 0.12 | Negative | Positive | 24.09 | No | N/A |
| 182 | 11.22 | Positive | 0.35 | Negative | 0.07 | Negative | Negative |  | Not reported | N/A |
| 191 | 7.87 | Negative | 1.75 | Positive | 8.41 | Positive | Negative |  | No | N/A |
| 213 | 140.62 | Positive | 0.00 | Negative | 0.11 | Negative | Negative |  | No | N/A |
| 223 | 7.29 | Negative | 4.73 | Positive | 7.35 | Positive | Negative |  | No | N/A |
| 228 | 0.17 | Negative | 5.56 | Positive | 40.35 | Positive | Negative |  | No | N/A |
| 232 | 16.48 | Positive | 0.19 | Negative | 0.09 | Negative | Positive | 23.89 | No | N/A |
| 246 | 5.60 | Negative | 1.37 | Positive | 6.98 | Positive | Negative |  | No | N/A |
| 255 | 8.06 | Negative | 1.09 | Positive | 10.18 | Positive | Negative |  | No | N/A |
| 280 | 20.61 | Positive | 2.30 | Positive | 0.51 | Negative | Positive | 32.34 | No | N/A |
| 296 | 15.70 | Positive | 0.04 | Negative | 0.15 | Negative | Negative |  | No | N/A |
| 319 | 4.07 | Negative | 2.48 | Positive | 1.16 | Positive | Negative |  | No | N/A |
| 328 | 6.41 | Negative | 0.68 | Negative | 2.59 | Positive | Negative |  | No | N/A |
| 337 | 80.22 | Positive | 0.86 | Negative | 5.62 | Positive | Negative |  | No | N/A |
| 355 | 2.36 | Negative | 2.11 | Positive | 0.10 | Negative | Positive | 29.18 | No | N/A |
| 357 | 10.49 | Positive | 3.34 | Positive | 0.73 | Negative | Negative |  | No | N/A |
| 360 | 13.57 | Positive | 0.06 | Negative | 0.09 | Negative | Negative |  | No | N/A |
| 376 | 45.70 | Positive | 0.07 | Negative | 0.09 | Negative | Negative |  | No | N/A |
| 387 | 310.78 | Positive | 0.46 | Negative | 4.56 | Positive | Positive | 24.24 | No | N/A |

Ct-cycle threshold; N/A-not applicable; PCR-polymerase chain reaction.
\*Mindray CL-900i anti-SARS-CoV-2 IgG assay positive: optical-density cutoff index $\geq 10.0$ vs. negative: cutoff index $< 10.0$ [6].
\*\*BioMérieux VidasIII assay positive: optical-density cutoff index $\geq 1.0$ vs. negative: cutoff index $< 1.0$ [7].
\*\*\*Roche Elecsys Anti SARS-CoV-2 assay positive: optical-density cutoff index $\geq 1.0$ vs. negative: cutoff index $< 1.0$ [5].
%Sample number consistent with Table S1 in supplementary information.

The overall, positive, and negative percent agreements between the CL-900i and the Elecsys assays were estimated at 93.4% (95% CI 90.5%-95.5%), 92.8% (95% CI 87.9%-95.8%), and 93.8% (95% CI 89.9%-96.3%), respectively (Table 2A). Cohen’s kappa statistic was estimated at 0.87 (95% CI 0.83-0.90), indicating excellent agreement between the two assays (Table 2A). The Spearman correlation between the optical densities was at 0.751 (p-value<0.001) indicating strong correlation.

**Table 2.** Concordance metrics between A) the Mindray CL-900i anti-SARS-CoV-2 IgG testing and the Roche Elecsys Anti SARS-CoV-2 testing, B) the VidasIII anti-SARS-CoV-2 IgG testing and the Roche Elecsys Anti SARS-CoV-2 testing, and C) the BioMérieux VidasIII anti-SARS-CoV-2 IgG testing and the Mindray CL-900i anti-SARS-CoV-2 IgG testing.

|  |  | Roche Elecsys<br>Anti SARS-CoV-2 |  |  | Overall<br>percent<br>agreement | Positive<br>percent<br>agreement | Negative<br>percent<br>agreement | Cohen's<br>kappa<br>statistic |
| --- | --- | --- | --- | --- | --- | --- | --- | --- |
|  |  | Positive | Negative | Total | % (95 CI) | % (95 CI) | % (95 CI) | k (95% CI) |
| **Mindray CL-900i**<br>**anti-SARS-CoV-2**<br>**IgG** | Positive | 155 | 14 | 169 | 93.4%<br>(90.5%-95.5%) | 92.8%<br>(87.9%-95.8%) | 93.8%<br>(89.9%-96.3%) | 0.87<br>(0.83-0.90) |
|  | Negative | 12 | 213 | 225 |  |  |  |  |
|  | Total | 167 | 227 | 394 |  |  |  |  |

\*CI-confidence interval

B)
|  |  | Roche Elecsys<br>Anti SARS-CoV-2 |  |  | Overall<br>percent<br>agreement | Positive<br>percent<br>agreement | Negative<br>percent<br>agreement | Cohen's<br>kappa<br>statistic |
| --- | --- | --- | --- | --- | --- | --- | --- | --- |
|  |  | Positive | Negative | Total | % (95 CI) | % (95 CI) | % (95 CI) | k (95% CI) |
| **BioMérieux**<br>**VidasIII anti-SARS-**<br>**CoV-2 IgG** | Positive | 155 | 5 | 160 | 95.7%<br>(93.2%-97.3%) | 92.8%<br>(87.9%-95.8%) | 97.8%<br>(95.0%-99.1%) | 0.91<br>(0.88-0.94) |
|  | Negative | 12 | 222 | 234 |  |  |  |  |
|  | Total | 167 | 227 | 394 |  |  |  |  |

\*CI-confidence interval

C)
|  |  | Mindray CL-900i<br>anti-SARS-CoV-2 IgG |  |  | Overall<br>percent<br>agreement | Positive<br>percent<br>agreement | Negative<br>percent<br>agreement | Cohen's<br>kappa<br>statistic |
| --- | --- | --- | --- | --- | --- | --- | --- | --- |
|  |  | Positive | Negative | Total | % (95 CI) | % (95 CI) | % (95 CI) | k (95% CI) |
| **BioMérieux**<br>**VidasIII anti-SARS-**<br>**CoV-2 IgG** | Positive | 151 | 9 | 160 | 93.2%<br>(90.2%-95.3%) | 89.3%<br>(83.8%-93.2%) | 96.0%<br>(92.6%-97.9%) | 0.86<br>(0.82-0.90) |
|  | Negative | 18 | 216 | 234 |  |  |  |  |
|  | Total | 169 | 225 | 394 |  |  |  |  |

\*CI-confidence interval.

The overall, positive, and negative percent agreements between the VidasIII and the Elecsys assays were estimated at 95.7% (95% CI 93.2%-97.3%), 92.8% (95% CI 87.9%-95.8%), and 97.8% (95% CI 95.0%-99.1%), respectively (Table 2B). Cohen’s kappa statistic was estimated at 0.91 (95% CI 0.88-0.94), indicating excellent agreement between the two assays (Table 2B). The Spearman correlation between the optical densities was at 0.824 (p-value<0.001) indicating strong correlation.

The overall, positive, and negative percent agreements between the VidasIII and the CL-900i assays were estimated at 93.2% (95% CI 90.2%-95.3%), 89.3% (95% CI 83.8%-93.2%), and 96.0% (95% CI 92.6%-97.9%), respectively (Table 2C). Cohen’s kappa statistic was estimated at 0.86 (95% CI 0.82-0.90), indicating excellent agreement between the two assays (Table 2C). The Spearman correlation between the optical densities was at 0.804 (p-value<0.001) indicating strong correlation.

A total of 49 swabs were PCR-positive at the time of specimen collection during this study for a current-infection prevalence of 12.5% (49/392; 95% CI 9.6%-16.1%)—two individuals declined PCR testing (but not antibody testing) during this study. Figure 1A shows the distribution of the PCR Ct values among those PCR-positive, indicating broad distribution suggestive of these persons being diagnosed at the various stages of infection. The mean PCR Ct value was 25.8 with a standard deviation of 6.3.

**Figure 1.**
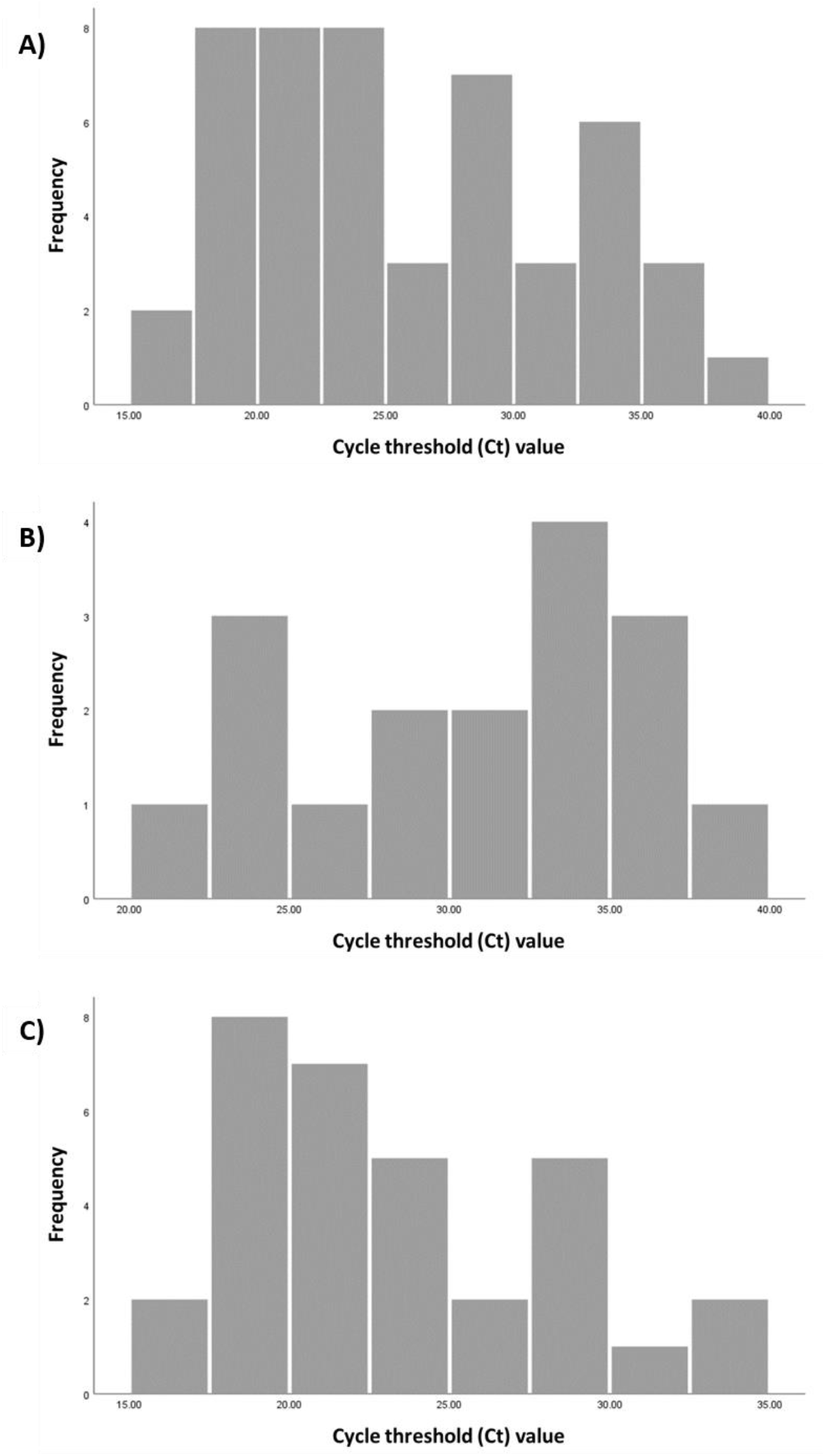
Distribution of polymerase chain reaction (PCR) cycle threshold (Ct) values of A) the 49 persons identified as SARS-CoV-2 PCR-positive at the time of specimen collection during the study, B) the 17 persons identified as SARS-CoV-2 PCR-positive, but were antibody-positive in at least one of the assays, and C) the 32 persons identified as SARS-CoV-2 PCR-positive, but were antibody-negative in all three assays.

Among all those PCR-positive at the time of specimen collection, 30.6% (15/49; 95% CI 19.5%-44.5%) were seropositive in the CL-900i assay, 22.5% (11/49; 95% CI 13.0%-35.9%) were seropositive in the VidasIII assay, 24.5% (12/49; 95% CI 14.6%-38.1%) were seropositive in the Elecsys assay, and 34.7% (17/49; 95% CI 22.9%-48.7%) were seropositive in at least one of the assays. Therefore, 32 individuals had a PCR-positive diagnosis at the time of specimen collection, but were antibody-negative in all three assays.

Among all those seropositive in at least one of the assays, 9.3% were PCR-positive (17/183; 95% CI 5.9%-14.4%) at the time of specimen collection. The mean PCR Ct value was 30.8 with a standard deviation of 5.2, indicative of mostly *non-recent* infections (Figure 1B). Among all those seronegative in all three assays, 15.2% were PCR-positive (32/211; 95% CI 11.0%-20.6%) at the time of specimen collection. The mean PCR Ct value was 23.0 with a standard deviation of 5.1, indicative of mostly *recent* infections (Figure 1C).

Through linking with the national SARS-CoV-2 PCR testing database [17], and of the 394 participants, 4.3% (17/394; 95% CI 2.7%-6.8%) had a record of SARS-CoV-2 PCR-confirmed diagnosis *prior* to this study. All but one of these were antibody-positive by at least one of the assays. The individual testing antibody-negative but had a prior PCR-confirmed diagnosis was diagnosed on July 23, 2020, that is four days prior to the antibody serological test date. This individual declined PCR testing during this study and at the time of the serological test.

Of the 183 persons with an antibody-positive status in at least one assay, 16 persons had a SARS-CoV-2 PCR-confirmed diagnosis *prior* to this study. Accordingly, the detection rate (the percentage of those antibody-positive who had a *prior* PCR-confirmed diagnosis) was 8.7% (16/183; 95% CI 5.5%-13.7%).

Based on the above, a total of 216 persons had a *laboratory-confirmed* infection at or prior to this study; that is an antibody-positive result in at least one assay (183 cases), a PCR-positive diagnosis prior to this study but with an antibody-negative status in all three assays (1 case), or a PCR-positive diagnosis at the time of specimen collection during this study but with an antibody-negative status in all three assays (32 cases). Accordingly, the percentage of persons with evidence of *ever infection* through either PCR or antibody testing was 54.8% (216/394; 95% CI 49.9%-59.7%). Moreover, only 78.2% of ever infections were antibody-positive by the CL-900i assay (169/216; 95% CI 72.3%-83.2%), 74.1% by the VidasIII assay (160/216; 95% CI 67.8%-79.5%), and 77.3% by the Elecsys assay (167/216; 95% CI 71.3%-82.4%).

Linking with the national COVID-19 hospitalization database [17] identified only one laboratory-confirmed infection through this study to have progressed to severe disease per WHO severity classification [18]. The person had also a diagnosis of diabetes, hypertension, and coronary artery disease. This person was diagnosed PCR-positive at time of specimen collection, was seronegative in the CL-900i and the VidasIII assays, but was seropositive in the Elecsys assay. No infection was critical per WHO severity classification [18] and no COVID-19 death was reported for any of the study participants.

## Discussion

A primary finding of this study is that all three antibody assays had comparable performance and excellent agreement. This positive finding, however, conceals important shortcomings about the use and performance of commercial antibody assays in assessing *ever infection* with SARS-CoV-2 in population-based surveys, especially at times of high SARS-CoV-2 incidence, as is the case at present globally.

The first shortcoming is that each of these three assays missed ≥69% of those who were PCR-positive at the time of specimen collection. This finding is explained in large part by the 1-4 weeks delay in development of detectable antibodies after acquiring the infection [20, 21]. This explanation is supported by the low PCR Ct value among those PCR-positive but antibody-negative (Figure 1C), which indicates recency of infection. At the time of the study and in the population being studied, the outbreak was advancing, so there was a significant proportion of new infections, making the serology assay less useful for estimating population prevalence of ever infection. It is unknown whether the lower sensitivity could have been due in part also to commercial assay development preferentially opting to maximize the specificity of the assay, to avoid a false positive diagnosis with its clinical implications, but at the expense of the sensitivity of the assay.

The second shortcoming is that each of these three assays also missed other individuals with evidence of ever infection. Despite excellent agreement overall, nearly 10% of the total sample still showed discordant results between the three antibody assays. Differences in the sensitivity of the assays to diagnose recent infection explains only partially these discordant results. Indeed, most (74.3%) of these persons with discordant results were *PCR-negative* at the time of specimen collection (Table 1), and thus less likely to have had a recent infection. The extent to which false positivity may explain some of these discordant results is unknown, but the three manufacturers reported essentially perfect specificity for each of these assays [5, 7, 22, 23].

As a consequence of these findings, the use of any one of these antibody assays to assess ever infection in a population-based sample, especially at the times of high SARS-CoV-2 incidence, will substantially underestimate the ever infection prevalence in the sample. In the sample in the present study, at least 20% of ever infections were missed. A solution to this challenge is to combine PCR data and serology data together, or that the serology data cannot be adequately interpreted without knowledge of the PCR positivity data, or that serology is less useful when the epidemiology is rapidly changing. With the global pandemic continuing at high SARS-CoV-2 incidence, this finding suggests that ever infection in populations is possibly substantially higher than is currently believed.

This study has some limitations. Two out of the 394 participants included in the study declined PCR testing (but not serological testing) at time of specimen collection. The performance of these antibody assays was compared to each other (and to PCR testing), but not to a gold standard test of seropositivity, as such a test was not available to study investigators. Therefore, we were unable to measure ever infection prevalence to a gold standard, and use this to compare the performance of each assay to the gold standard, nor to assess the sensitivity and specificity of each assay in the study sample. The specificity of the Elecsys assay has previously been reported to be 99.98% and the sensitivity to be 98.80% on day 14 after PCR diagnosis [5]. A validation study by Public Health England reported a specificity of 100% and a sensitivity of 83.9% for the same assay [22]. As for the remaining assays, specificity and sensitivity were reported at 94.9% and 82.2%, respectively, for the CL-900i assay [23], and at 99.9% and 88.6%, respectively, for the VidasIII assay [7].

In conclusion, all three assays had comparable performance and excellent agreement when used in a high SARS-CoV-2 exposure setting, but still missed at least 20% of cases with laboratory-confirmed evidence of ever infection. This suggests that current growing use of commercial antibody assays to assess ever infection in population-based surveys, especially at times of high SARS-CoV-2 incidence when many infections are recent, is likely to substantially underestimate actual infection exposure. The findings demonstrate further the need to interpret the serology testing together with PCR testing.

## Data Availability

All relevant data are available within the manuscript.

## Acknowledgments

We thank Her Excellency Dr. Hanan Al Kuwari, Minister of Public Health, for her vision, guidance, leadership, and support. We also thank Dr. Saad Al Kaabi, Chair of the System Wide Incident Command and Control (SWICC) Committee for the COVID-19 national healthcare response, for his leadership and analytical insights, and for his instrumental role in enacting data information systems that made these studies possible. We further extend our appreciation to the SWICC Committee and the Scientific Reference and Research Taskforce (SRRT) members for their informative input, scientific technical advice, and enriching discussions. We also thank Dr. Mariam Abdulmalik, CEO of the Primary Health Care Corporation and the Chairperson of the Tactical Community Command Group on COVID-19, as well as members of this committee, for providing support to the teams that worked on the field surveillance. We further thank Dr. Nahla Afifi, Director of Qatar Biobank (QBB), Ms. Tasneem Al-Hamad, Ms. Eiman Al-Khayat and the rest of the QBB team for their unwavering support in retrieving and analyzing samples and in compiling and generating databases for COVID-19 infection, as well as Dr. Asmaa Al-Thani, Chairperson of the Qatar Genome Programme Committee and Board Vice Chairperson of QBB, for her leadership of this effort. We also acknowledge the dedicated efforts of the Clinical Coding Team and the COVID-19 Mortality Review Team, both at Hamad Medical Corporation, and the Surveillance Team at the Ministry of Public Health. We thank Dr. Katharine J. Looker, medical writer from the University of Bristol, for reviewing the manuscript.

## Author contributions

AAT conceived the study. LJA, GKN, and SRD co-designed the study and led the statistical analyses. GKN and PC led the laboratory testing. FS, DWA, and HA performed the CL-900i and the VidasIII testing. SRD, HC, and LJA performed the data analyses and wrote the first draft of the article. All authors contributed to data collection and acquisition, database development, discussion and interpretation of the results, and to the writing of the manuscript. All authors have read and approved the final manuscript.

## Data and materials availability

All relevant data are available within the manuscript.

